# Impostor Phenomenon, Psychological Capital, and Psychological Distress among Pakistani University Students: A Cross-Sectional Study

**DOI:** 10.1101/2025.09.09.25334787

**Authors:** Roomana Pervaiz, Tamkeen Ashraf Malik

## Abstract

**Background:** Impostor Phenomenon (IP) is linked with adverse mental health outcomes among university students whereas Psychological Capital (PsyCap) may work as a personal resource linked with improved mental health outcomes. This study examines the relationship of IP and PsyCap with Psychological Distress (PD) among university students and if this association differs among male and female students.

**Method:** A cross sectional study was conducted among university students (N = 323) who completed measures of IP, PsyCap and PD. Pearson correlation was used to measure the association among the mentioned variables. Hierarchical multiple regression was conducted to assess the predictive effect of IP and PsyCap on PD after controlling for gender, educational group, and recent stressor. Exploratory gender analysis and a separate regression analysis for depression, anxiety and stress ware also conducted.

**Results:** PD was positively correlated with IP (r =.595, p <.01) and negatively associated with PsyCap (r = -.282, p <.01). In the hierarchical regression, the addition of IP and PD explained an additional variance of 37% in PD (ΔR² =.37, p <.001), increasing the total variance to 42.2% (R² =.422). IP emerged as the strongest predictor (β =.562, p <.001), and PsyCap showed significant negative predictive effect (β =-.147, p <.001). Exploratory gender analysis revealed that IP significantly predicted PD among both male (β =.587, p <.001) and female students (β =.554, p <.001) while PsyCap demonstrated significant negative association with PD among females (β =-.156, p <.004), not males (β =.141, p <.130). Regression analysis of dimensions of PD indicated that IP significantly predicted stress (β =.532), anxiety (β =.499) and depression (β =.507), all p <.001.

**Conclusion:** The findings indicate that IP is strongly associated with PD among university students and have similar predictive effect on male and female students. PsyCap showed a modest negative association with PD. The study highlights the importance of impostor experiences for university students and cultivating psychological resources to support mental health and wellbeing of university students.

---

Impostor Phenomenon, Psychological Capital, and Psychological Distress among Pakistani University Students: A Cross-Sectional Study Psychological distress (PD), in this research, refers to the combined symptoms of depression, anxiety and stress. These symptoms emerge when an individual’s coping ability is insufficient to deal with stressful demands. Global data show that young adults report disproportionately higher levels of mental health problems, a trend exacerbated during and after COVID-19 (WHO, 2022). Psychological distress has been associated with adverse outcomes such as suicidal behaviour, academic difficulties and physical health consequences, academic difficulties (Chaudhry et al 2026), as well as a range of adverse physical health outcomes (Harvanek et al., 2021). Therefore, it is imperative to understand the factors contributing to psychological distress.

A recent study among Pakistani psychology students also reported substantial levels of depression (51.6%), anxiety (62%), and stress (41.4%), highlighting the mental health vulnerability of university students in Pakistan (Siddiqui et al., 2026). University students may be vulnerable because of academic, social and developmental demands of their age. Reported distress prevalence in Pakistan ranges from 35% to 85% (Ullah et al., 2022; Asif et al., 2020) with a higher prevalence among females (Viertio, et al, 2021; Bor et al., 2018). Factors such as academic pressure, societal expectations, and uncertainty about careers can increase vulnerability to distress.

University students may experience self-doubt, low self-esteem and fear of failure - features of impostor phenomenon (IP) (Siddiqui, Hamayunm & Khan, 2022) which is characterized by persistent self-doubt regarding their abilities and tendency to internalize failures and externalize achievements (Clance & Imes, 1978). Cognitive theory suggests that people with IP may have dysfunctional beliefs that they are intellectually not enough despite the counter evidence. These maladaptive cognitions may lead to negative automatic thoughts, increased self-criticism, and fear of failure, thus increasing vulnerability to depression, anxiety and stress. According to the literature, prevalence of IP ranges from 9% to 89% (Bravata et al., 2020). Moreover, intense IP experiences are correlated with higher levels of stress (Carlos et al., 2023), anxiety (El-Ashry et al., 2024), and depression (Qasem et al., 2025). Thus, IP emerges as a potential correlate of PD that warrants further investigation

The data is inconsistent regarding gender difference in IP. A systematic analysis reported that nearly half of the studies under the review reported no gender differences, whereas other studies reported higher levels of IP among women (Bravata et al., 2020; Jansson et al., 2025; Qureshi et al., 2025). Given these inconsistent findings, gender was added as a covariate in the present study. In addition to gender, level of education is also associated with IP. Evidence also suggests difference in IP across educational levels; therefore, educational status was also included as a covariate in the present study.

Conservation of resources theory proposes that individuals tend to obtain, retain and protect valued personal resources. PD is experienced when these personal resources are threatened. PsyCap is an higher order construct that consists of hope, self-efficacy, resilience, and optimism (abbreviated as HERO). It is an crucial personal resource that helps individuals deal with the stressors in life. Existing researches reveal that it is positively correlated with academic success (Hu et al., 2025) and negatively linked with PD (Chan et al., 2026; Quinde et al., 2026; Pradhan et al., 2021). Therefore, higher PsyCap may serve as an important personal resource linked with lower levels of PD among university students.

However, research on IP remains concentrated in specific population, especially medical trainees (Tahir et al., 2020) with limited research on broader university population.

Furthermore, limited research has examined IP, PsyCap and PD simultaneously in Pakistani university students. Therefore, examining these factors is imperative to obtain a comprehensive understanding of PD. The present study aimed to investigate the relationships among IP, PsyCap and PD in Pakistani university students. It also examined whether IP and PsyCap significantly predicted PD after controlling for gender, educational status, and recent stressful experience. Based on the literature reviewed, the following hypothesis were proposed:

H1. Impostor phenomenon will be positively associated with psychological distress, whereas psychological capital will be negatively associated with psychological distress among university students.

H2. Impostor phenomenon and psychological capital will significantly predict psychological distress after controlling for gender, educational status, and recent stressful experience.

## Methodology

The research adopts a correlational quantitative approach using a cross-sectional survey design. Non-probability convenient sampling method was used to recruit participants in 2021 from coeducational universities in Islamabad, mainly the National University of Sciences and Technology (NUST) and Quaid-e-Azam University (QAU). Both paper-based and online questionnaires (through Google Forms) were used to recruit students through university cafeterias, hostels, and personal networks.

Eligible participants were undergraduate or postgraduate students aged 18-34 who could read Urdu or English and provide informed consent. Written and electronic consent was provided. For in-person recruitment, the researcher also provided verbal clarification and answered questions when required. Study information was provided at the beginning of both versions of the questionnaire. Participation was voluntary, and a random-draw incentive was offered; however, the selected winners did not respond and therefore no incentive was ultimately received. Incomplete responses and responses from individuals reporting current psychosis or inability to provide informed consent were excluded. A total of 323 participants were included in the final sample.

### Instruments

All instruments were self-report questionnaires administered on paper or online in English or Urdu as preferred by the participants.

**Clance Impostor Phenomenon Scale (CIPS)** was used to measure IP (Clance, 1985). It is a 5-point likert scale consisting of 20 items with high consistency (a=0.85-0.96) and proven reliability in identifying impostors from non-impostors (Mak et al., 2019). It has four categories (few/mild, moderate, frequent, and intense) with high scores indicating more reported symptoms. For correlational analysis, the total score was treated as continuous variable. In the present sample, the CIPS demonstrated good internal consistency (Cronbach’s α =.86).

### Psychological Capital Questionnaire (PCQ-12)

Psychological Capital Questionnaire (PCQ-12; Luthans et al., 2007) is a 12-item scale whose Urdu version (Abbasi, 2015) was used to assess PsyCap in this study. It measures four dimension of PsyCap (HERO) on a six-point likert scale. Higher scores indicate higher levels of PsyCap. Urdu version was used in the study. The PCQ-12 demonstrated good internal consistency in the present sample (Cronbach’s α =.81).

### Depression, Anxiety and Stress Scale-21 (DASS-21)

Symptoms of depression, anxiety and stress were measured using DASS-21 (Lovibond & Lovibond, 1995). This self-report measure contains 21-items which are rated on a four-point scale from 0 to 3 based on symptoms experience by the participant during last one week. For the present study, psychological distress was operationalized as a composite score representing overall symptom burden across the three DASS dimensions. The 21 items were summed and multiplied by 2 to obtain scores comparable with the DASS-42 scoring metric, with higher scores indicating greater symptom burden. Depression, anxiety, and stress subscale scores were also examined in secondary analyses. An established Urdu version of the DASS-21 was used where preferred by participants. The scale showed an excellent internal consistency in the present sample (Cronbach’s α =.94).

### Statistical Analysis

After data entry and data screening, statistical analysis was conducted using SPSS Windows version 19. Pearson product-moment correlation coefficient was used to assess bivariate correlations. Hierarchical multiple regression was used to examine whether IP and PsyCap predicted PD after controlling for gender, educational status, and recent stressful experience. The data met assumptions of homoscedasticity and multicollinearity were examined. Finally, in the regression analysis, IP and PsyCap was modeled as predictor, PD as dependent variable as the outcome and experience of recent stressor, gender, educational status as covariates. Statistical significance was defined as p <.005. Gender stratified models were run to account for possible gender variations in the correlations among the variables.

## Results

The study sample consisted of 239 female (74%) and 84 males university students (26.0%) in the age ranging from 18 to 34 years (*M* = 24.57, *SD* = 2.60). Age, nevertheless, was available for 296 participants. Undergraduate university students make up 48.3% (n = 145) of the study sample whereas 51.7% (n = 155) were postgraduate students. Most participants (70.0%) reported no significant stressful experience in the past six months.

**Table 1.** Demographic characteristics of both male and female university students (N=323).

| Characteristic | *n* | % |
| --- | --- | --- |
| **Gender** |  |  |
| Male | 84 | 26.0 |
| Female | 239 | 74.0 |
| **Age, years** |  |  |
| *M* (SD) | 24.57 (2.60) |  |
| Range | 18–34 |  |
| Missing | 27 |  |
| **Educational status** |  |  |
| Undergraduate | 145 | 48.3 |
| Postgraduate | 155 | 51.7 |
| Missing | 23 |  |
| **Recent stressful experience** |  |  |
| Yes | 96 | 30.0 |
| No | 224 | 70.0 |
| Missing | 3 |  |

**Note.** IP = impostor phenomenon; PD = psychological distress; PsyCap = psychological capital; RSE = recent stressful experience. Percentages for educational status and RSE are based on valid responses.

**Table 2.**

| Variable | *M* | *SD* | $\alpha$ | 1 | 2 | 3 |
| --- | --- | --- | --- | --- | --- | --- |
| 1. Impostor Phenomenon (CIPS) | 59.51 | 13.57 | .86 |  | -.191** | .595** |
| 2. Psychological Capital | 4.23 | .71 | .81 |  | — | -.282** |
| 3. Psychological Distress | 45.90 | 27.93 | .94 |  | — | — |

*Note.* *N* = 323. CIPS = Clance Impostor Phenomenon Scale; $\alpha$ = Cronbach's alpha. **P** < .01. Impostor phenomenon was positively associated with psychological distress, whereas psychological capital was negatively associated with distress. Impostor phenomenon and psychological capital were also negatively associated with each other.

### Hierarchical Multiple Regression

A hierarchical multiple regression analysis was conducted to investigate the predictive effect of IP and PsyCap on PD after accounting for demographic characteristics. 298 complete cases were included in the regression analysis. Twenty-five participants did not respond to one or more of the variables included in the regression analysis; thus, those cases were excluded. Gender, educational status and recent stressful experience were entered at initial step followed by impostor phenomenon and psychological capital at the second step.

**Table 3.** Hierarchical Multiple Regression Predicting Psychological Distress. (N=298).

| Predictor | B | SE B | $\beta$ | t | p | 95% CI for B |
| --- | --- | --- | --- | --- | --- | --- |
| **Step 1** |  |  |  |  |  |  |
| Gender | 2.237 | 3.703 | .036 | 0.604 | .546 | (-5.052, 9.525) |
| Educational status | 6.350 | 3.280 | .114 | 1.936 | .054 | (-0.106, 12.806) |
| Recent stressful experience | -11.142 | 3.478 | -.182 | -3.203 | .002 | (-17.987, -4.296) |
| **Step 2** |  |  |  |  |  |  |
| Gender | -0.891 | 2.911 | -.014 | -0.306 | .760 | (-6.620, 4.839) |
| Educational status | 6.833 | 2.577 | .122 | 2.652 | .008 | (1.762, 11.904) |
| Recent stressful experience | -9.416 | 2.729 | -.154 | -3.450 | .001 | (-14.787, -4.045) |
| Impostor phenomenon | 1.153 | 0.094 | .562 | 12.263 | < .001 | (0.968, 1.338) |
| Psychological capital | -5.751 | 1.788 | -.147 | -3.216 | .001 | (-9.271, -2.232) |

**Note.** N = 298. Gender, educational status, and recent stressful experience were entered at Step 1 ( $R^2 = .052$ , adjusted $R^2 = .042$ , $F(3, 294) = 5.38$ , $p < .001$ ); IP and psychological capital were entered at Step 2 ( $R^2 = .422$ , adjusted $R^2 = .412$ , $\Delta R^2 = .370$ , $\Delta F(2, 292) = 93.45$ , $p < .001$ ). Recent stressful experience was coded 1 = yes and 2 = no.

### Exploratory Gender-Stratified Analyses

**Table 4.** Gender-stratified hierarchical regression analysis predicting Psychological Distress among university students (N=298).

| Predictor | Male (n = 80) |  | Female (n = 218) |  |
| --- | --- | --- | --- | --- |
| | $\beta$ | p | $\beta$ | p |
| **Step 1** |  |  |  |  |
| Educational Status | .160 | .081 | .106 | .043 |
| Recent Stressful Experience | -.132 | .148 | -.162 | .002 |
| **Step 2** |  |  |  |  |
| IP | .587 | < .001 | .554 | < .001 |
| PsyCap | -.141 | .130 | -.156 | .004 |
| **Model Summary** |  |  |  |  |
| Step 1 R<sup>2</sup> (Adj R<sup>2</sup>) | .031 (.006) |  | .053 (.045) |  |
| Step 2 R<sup>2</sup> (Adj R<sup>2</sup>) | .420 (.389) |  | .421 ( .410) |  |
| $\Delta R$ | .389 | | .368 | |
| $\Delta F$ | 25.12* | | 67.65* | |

*Note: For $\Delta F$ values, both models were significant ( $p < .001$ )*

Exploratory gender stratified analysis reveals that the addition of IP and PsyCap significantly improved model fit for both genders included in the study for both male (Change R =.389, p = <.001) and female students.

**Table 5.**
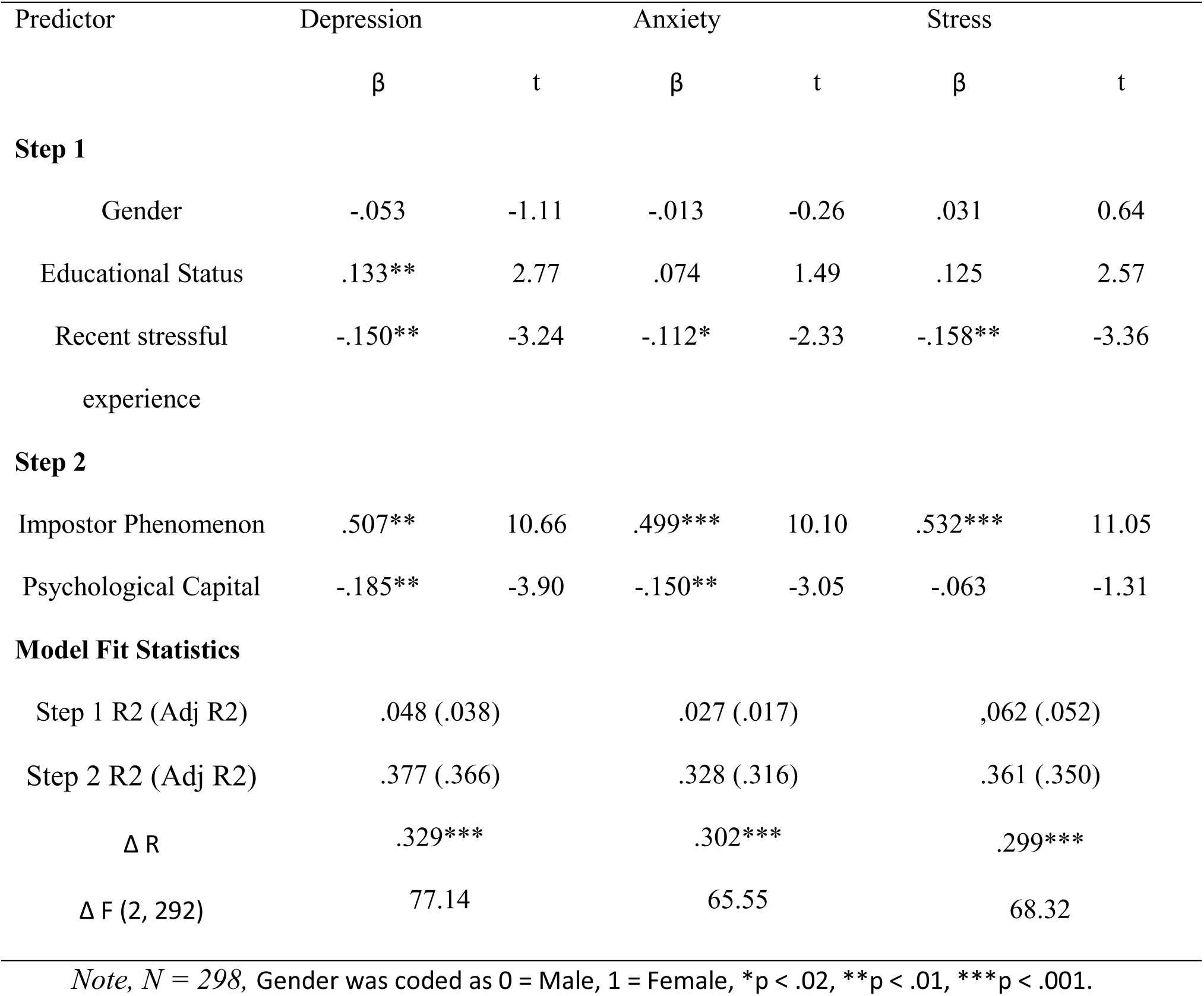
Exploratory hierarchical regression analyses predicting depression, anxiety, and stress (N=298).

| Predictor | Depression |  | Anxiety |  | Stress |  |
| --- | --- | --- | --- | --- | --- | --- |
| | $\beta$ | t | $\beta$ | t | $\beta$ | t |
| **Step 1** |  |  |  |  |  |  |
| Gender | -.053 | -1.11 | -.013 | -0.26 | .031 | 0.64 |
| Educational Status | .133** | 2.77 | .074 | 1.49 | .125 | 2.57 |
| Recent stressful<br>experience | -.150** | -3.24 | -.112* | -2.33 | -.158** | -3.36 |
| **Step 2** |  |  |  |  |  |  |
| Impostor Phenomenon | .507** | 10.66 | .499*** | 10.10 | .532*** | 11.05 |
| Psychological Capital | -.185** | -3.90 | -.150** | -3.05 | -.063 | -1.31 |
| **Model Fit Statistics** |  |  |  |  |  |  |
| Step 1 R<sup>2</sup> (Adj R<sup>2</sup>) | .048 (.038) |  | .027 (.017) |  | .062 (.052) |  |
| Step 2 R<sup>2</sup> (Adj R<sup>2</sup>) | .377 (.366) |  | .328 (.316) |  | .361 (.350) |  |
| $\Delta R$ | .329*** | | .302*** | | .299*** | |
| $\Delta F$ (2, 292) | 77.14 | | 65.55 | | 68.32 | |

*Note, N = 298, Gender was coded as 0 = Male, 1 = Female, \*p < .02, \*\*p < .01, \*\*\*p < .001.*

Demographic variables were entered at Step 1 followed by addition of IP and PsyCap at Step 2. Δ*R*² represents the additional variance explained by CIPS and psychological capital at Step 2. For the final models, *R*² =.377 for depression,.328 for anxiety, and.361 for stress.

Although previous literature has produced inconsistent findings regarding gender differences in impostor phenomenon, the association between impostor phenomenon and psychological distress was evident in both male and female students.

## Discussion

The present study aimed to investigate the relationship among IP, PsyCap and PD among university students in Pakistan. In line with the proposed conceptual framework, PD was positively and negatively associated with IP and PsyCap respectively. Hierarchical regression further demonstrated that impostor phenomenon and psychological capital explained substantial additional variance in psychological distress beyond gender, educational status and recent stressful experience. Exploratory analyses showed that the positive association between impostor phenomenon and distress was consistent across depression, anxiety, and stress, whereas the protective association of psychological capital was evident for depression and anxiety but not stress.

One of the most significant findings of the present study was the positive correlation of IP with PD and its three dimensions. Students reporting intense impostor feelings also reported higher PD. This is in line with the existing literature linking IP with stress, anxiety, depression, burnout and emotional exhaustion among university students and early career professionals (Fleischhauer et al., 2023; Muradoglu et al., 2022; Bravata et al., 2020; Pateet et al., 2014). IP emerged as a major predictor of distress after accounting for gender, educational group and recent stressful experience. Cognitive theory opines that people with IP may interpret academic difficulties as proof of failure and personal inadequacy while ignoring the evidence that suggests otherwise. These dysfunctional thoughts may invite self-doubt, self-criticism and negative automatic thoughts which in turn may lead to distress. Therefore, the findings of the study provide empirical support for the hypothesized cognitive pathway associating IP with PD.

As far as PsyCap is concerned, it demonstrated negative correlation with PD which suggests the link between personal psychological resources and better psychological functioning. This finding is corroborated by findings of Pradhan et al. (2021), Avey et al. (2011) and Newman et al. (2014). Conservation of resources theory suggests that psychological capital may serve as a personal resource that helps in dealing with academic demands, and alleviates distress. Nevertheless, further exploratory analysis shows that the predictive effect of PsyCap was not consistent across all DASS dimension; it predicted anxiety and depression, not stress after controlling demographic variables and IP. A tentative explanation could be that stress seems to me more affected by the academic demands as compared to relatively stable personal psychological resources. However, this explanation needs further investigation in future through longitudinal research.

The results revealed that gender did not predict distress significantly after controlling for educational status, IP and PsyCap. IP and PD were related to each other in both male and female students. This is corroborated by existing research that suggests that IP is not limited to females only. The uniformity of IP coefficients across males and females demonstrates that mental processes involved in IP are relevant to students irrespective of gender. Nevertheless, it is imperative to mention that the gender-stratified analysis were not the formal tests of moderation; they were exploratory. Thus, it cannot be established that gender has a role in altering the strength of these associations. The findings also reveal that educational status was associated with PD which means that academic demands vary across educational groups.

Previous researches on IP are mostly focused on western population. This study extends the emerging literature on IP beyond the Western samples. It also provides evidence that the association between IP and PD is relevant within a South Asian context. Academic competition, career uncertainty, family expectations, and culturally shaped perceptions of achievement may provide important contextual conditions in which impostor experiences are expressed. These mechanisms, however, were not directly assessed in the present study.

The findings of the study have implication for university mental health as well as for institutions responsible for mental health of students. Students should be screened for maladaptive and dysfunctional thought patterns, perfectionistic tendencies and other impostor related experiences for early identification and assessment followed by psycho-educational programs. These programs can help cultivate psychological resources in addition to targeting other interventions.

## Limitations

Since the study used cross sectional research design, causality cannot be established. Future researchers can adopt longitudinal design on various Pakistani universities to examine the minimizing impact of PsyCap on the association between IP and PD. Also, convenient sampling limits generalizability. Furthermore, the study has more female participants which limit the accuracy of estimates for male students. Future research can examine the moderating effect of PsyCap on the relationship between IP and PD.

## Conclusion

The study provides empirical evidence that IP is a significant correlate of psychological distress among university students especially among university students. Intense IP experiences were related to higher distress, depression, anxiety, and stress. Psychological capital was linked with lower overall distress, depression, and anxiety. The findings highlight the importance of addressing maladaptive self-evaluations while strengthening psychological resources within university mental health initiatives.

Longitudinal research is needed to clarify the directionality of these relationships and determine whether enhancing psychological capital can reduce vulnerability to distress associated with impostor experiences.

## Data Availability

All data produced in the present study are available upon reasonable request to the authors.

